# Adapting reactive case detection strategies for high-risk populations to increase malaria infection detection in Aceh Province, Indonesia

**DOI:** 10.1101/2025.10.22.25337765

**Authors:** Adam Bennett, Chris Cotter, Iska Zarlinda, Martha G Silaen, Rosa Nora Lina, Carmen Cueto, Iqbal Elyazar, Jerry O. Jacobson, Lenny Ekawati, Michelle S Hsiang, Rintis Noviyanti, Jennifer L Smith, Farah N Coutrier

**Affiliations:** Malaria Elimination Initiative, Institute for Global Health Sciences, University of California San Francisco, San Francisco, California, USA; Department of Epidemiology and Biostatistics, University of California San Francisco, San Francisco, California, USA; Department of Women’s and Children’s Health, Uppsala University, Uppsala, Sweden; Eijkman Research Center for Molecular Biology, National Research and Innovation Agency (BRIN); Jakarta, Indonesia; Oxford University Clinical Research Unit (OUCRU), Jakarta, Indonesia; Department of Pediatrics, University of California San Francisco, Benioff Children’s Hospital, San Francisco, California, USA

**Keywords:** malaria elimination, reactive case detection, high-risk populations, Aceh Province, Indonesia

## Abstract

**Introduction:** Reactive case detection (RACD), or testing and treatment of close contacts of recent malaria cases, is widely used in settings nearing malaria elimination, but often results in very low yield and uncertain impact. In areas where the primary vector exposure occurs outside the home, adapting RACD to target high-risk populations with specific exposure profiles may increase yield, surveillance value, and impact on treatment rates and transmission.

**Methods:** We evaluated the feasibility and yield of a risk-based RACD approach based upon forest-based exposure, in comparison with the standard neighborhood and household-based RACD conducted in Aceh Province, Indonesia. Index malaria cases were enrolled from public health facilities in five subdistricts. For each index case, standard RACD included testing members of households within 500 m, while risk-based RACD involved testing non-household contacts sharing a forest worksite or forest travel. Infection prevalence was measured by light microscopy, loop-mediated isothermal amplification (LAMP) and polymerase chain reaction (PCR).

**Results:** The study enrolled 47 index malaria cases during the study period, the majority (29) of which were diagnosed as *P. knowlesi* infections. Forty-one of the cases reported potential forest exposure outside the home, and follow-up was completed for 40 cases. Standard household-based RACD tested 847 household members from 34 index cases, yielding only 1 infection by LAMP/PCR (0.1%). Risk-based RACD tested 179 eligible contacts from 32 index cases, yielding 8 LAMP positive infections (4.5%), including 3 confirmed by PCR (1.7%).

**Conclusion:** In Aceh Province, targeting RACD to non-household contacts with shared high-risk exposures substantially increased infection yield compared to household-based RACD. Risk-based, exposure-driven approaches may improve the effectiveness of RACD in settings where malaria risk is greater outside the home.

## BACKGROUND

Substantial progress has been made recently in reducing malaria transmission in many countries to the point where elimination has been or can soon be achieved. While this progress is critical (1), malaria-eliminating countries face new technical and operational challenges to surveillance and response as malaria transmission declines (2). Traditionally, malaria control programs rely upon passive surveillance systems to monitor disease burden. In malaria elimination settings, however, the World Health Organization (WHO) recommends incorporating active surveillance methods such as case investigation followed by reactive case detection (RACD) to identify and clear additional infections that occur near index cases in space and time (3)(4). Visiting an index patient’s household and testing household members and neighbors allows programs to identify infection hot-spots, quickly treat infections, and implement foci management to prevent ongoing transmission (5).

In settings where transmission primarily occurs outside the household, such as through travel or occupational exposure, household-based RACD may yield few infections (6). In the Greater Mekong Subregion (GMS), where most exposure is thought to occur through forest or other work outside the home, observational studies have found limited benefit of household-based RACD (6). Across SE Asia, forest-working is a well-documented risk factor for malaria infection. In Aceh Province, Indonesia, (7) a previous study showed that when transmission occurs in the forest or other non-domestic settings, populations at higher risk of infection are likely to be linked through shared behaviors that increase their exposure to outdoor-biting and forest-dwelling mosquitos, such as sleeping outdoors or traveling and working together in forest or forest fringe areas.

Evidence remains limited on when implementing RACD targeted to high-risk populations is feasible and more effective than traditional household-based RACD as part of an elimination strategy. A study in Cambodia found that including individuals occupationally co-exposed with malaria index cases increased the test positivity rate from 0.16% to 3.9% (8). Another Cambodian study successfully identified more malaria infections by conducting RACD among co-travellers of index cases (9). Studies elsewhere have shown the potential benefit of targeting occupational contacts of malaria index cases, as well as the limited yield of household-based screening in areas with high forest exposure (10)(11).

In this study, we conducted a prospective malaria surveillance study in Aceh Province, Indonesia, a low transmission setting with a high proportion of *Plasmodium knowlesi* infections and predominantly forest-based livelihoods, to determine the feasibility of a risk-based RACD strategy. We compared characteristics of target populations and yield of secondary infections identified through risk-based RACD approaches to that of standard household-based RACD.

## MATERIALS AND METHODS

### Study design

A prospective surveillance study was conducted through the passive and reactive case detection program in Aceh Province, Indonesia between March 2017 and August 2018.

### Study site

Indonesia aims to achieve national elimination by 2030 through a spatially-progressive approach, eliminating malaria island-by-island. By 2017, over 70% of its population lived in malaria-free areas, and as of 2025, 79% of its administrative areas had achieved subnational elimination (12)(13). Following the 2009 National Ministerial Decree on Malaria Elimination, the Provincial Parliament of Aceh endorsed a malaria elimination target by 2020 (14). This goal was supported by strong political and financial support from the Government of Aceh, and Aceh Besar and Aceh Jaya successfully achieved elimination of non-zoonotic malaria in 2022 and 2025, respectively. Elimination strategies focused on improved passive and active surveillance, indoor residual spraying, long-lasting insecticide-treated bed net distribution, larvaciding, and environmental management. Since 2010, the Aceh Provincial Health Office included case investigation and RACD as part of routine malaria surveillance activities. This policy involves microscopy testing of index case household members and neighbors living within a 500 meter radius of each index case.

Aceh Province is a low-endemic transmission setting located on Sumatra Island in western Indonesia. Study activities were conducted in three subdistricts of Aceh Besar district (Kuta Cot Gli, Lembah Seulawah (Saree), and Lhoong) and two subdistricts of Aceh Jaya district (Krueng Sabe, Sampoiniet) (**Figure 1**). Each subdistrict has a single public health facility (puskesmas). These sites were selected as they represented the highest burden subdistricts and/or were part of a previous RACD study (7). Malaria transmission season in Aceh Province typically occurs from January to July. In 2017, there were 135 cases in Aceh Jaya out of a population of 87,622, for an annual parasite incidence (API) of 1.54, and 30 cases in Aceh Besar out of a population of 409,109 for an API of 0.07. The majority of cases in Aceh Jaya were *Plasmodium vivax* infections (93%), while the majority in Aceh Besar were *P. knowlesi* (80%).

**Figure 1:**
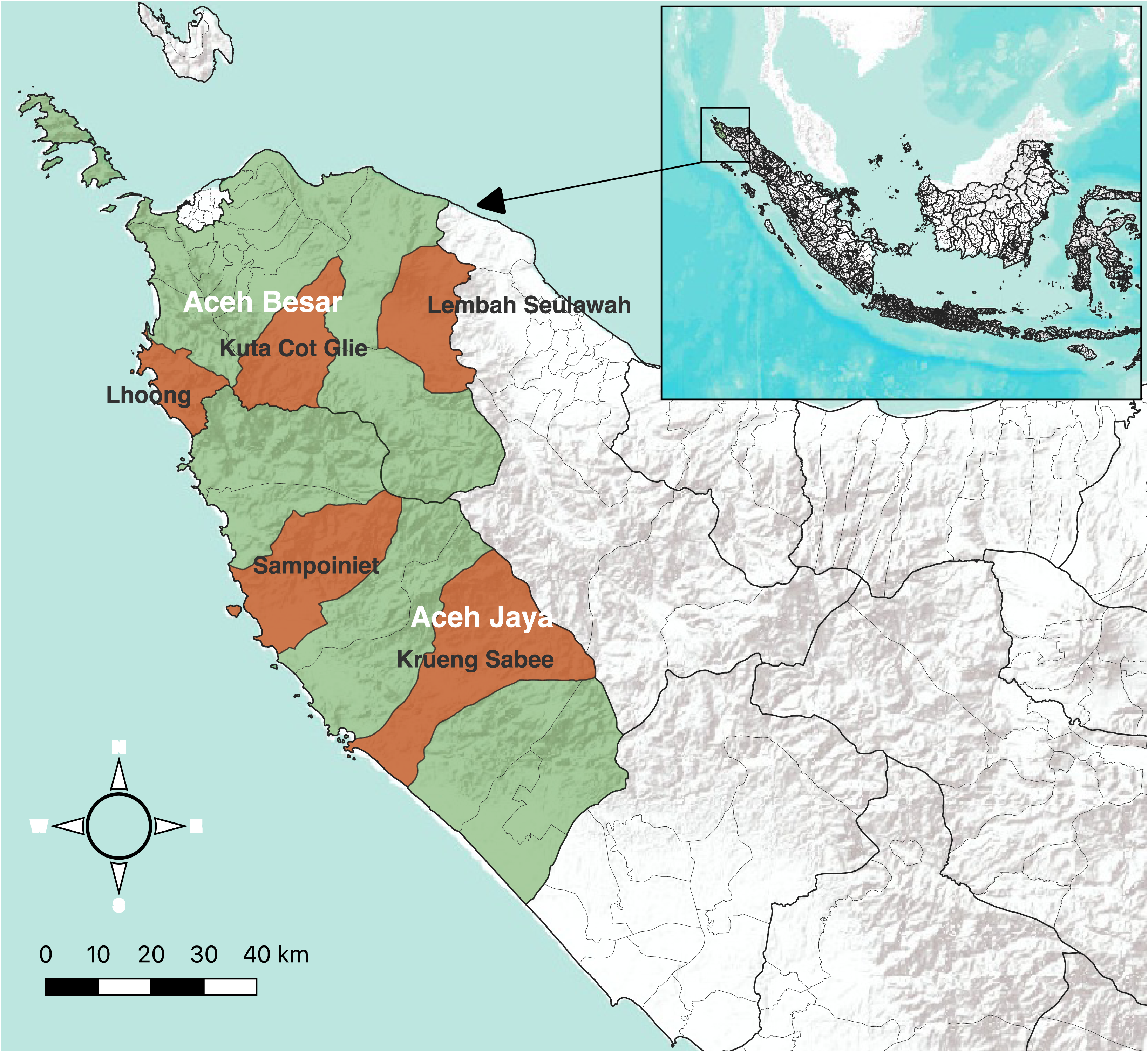
Selected study sub-districts in Aceh Besar and Aceh Jaya.

### Study population

A formative assessment in 2016 among forest workers and key informants in Aceh Besar and Aceh Jaya districts explored malaria risks associated with forest work and strategies for targeted surveillance, including ways to adapt RACD for increased infection detection (15). The present study population included all microscopy or LAMP confirmed cases of malaria at participating health facilities with a local residence and/or recent forest work in the district, household members and neighbors living within 500 meters of the index case, and for cases who reported forest or forest fringe work in the previous 60 days, individuals who worked together with the index case in or near the forest. For the eligibility criteria, ‘work’ was defined as paid activity or production/extraction of materials (e.g., gold, teak wood, fruits, vegetables, animals) primarily for sale as opposed to personal use.

### Health facility and field procedures

Patients presenting with suspected malaria at participating facilities were asked to provide a finger-prick blood sample for thick and thin smears and LAMP testing. Microscopy-positive patients meeting eligibility criteria were invited to participate. Microscopy-negative patients who tested positive by LAMP as part of a contemporaneous case-control study were also invited to participate (16). Written informed consent was obtained for the collection of an additional 250uL – 3 mL of blood by venous draw (age-dependent volume) prior to treatment, which was used to generate dried blood spots (DBS) and stored in heparin or EDTA tubes for molecular analysis. A brief index case questionnaire was administered to capture index case contact information and details on forest co-workers and work-sites (including employer contact information) visited overnight in the past 60 days.

Enrollment of a malaria case with forest exposure triggered one of two risk-based RACD responses, based on predefined criteria related to malaria risk, location, accessibility, and worksite size in the past 60 days. Venue-based RACD (VB-RACD) was conducted at eligible worksites that: (i) were located within the study district; (ii) were currently active or expected to be active within the next 60 days; (iii) employed six or more workers at a time; and (iv) were confirmed by the employer (*toke*) to be safe and accessible. Eligible sites were ranked by malaria risk and visited in priority order. At each site, field teams carried out community sensitization followed by all study activities—participant enrollment, questionnaire administration, and blood sample collection—in a private area. Peer referral-based RACD (PR-RACD) was used when worksites could not be visited in-person and/or employed fewer than six workers at one time. In this approach, the index case provided contact information for all co-workers and co-travelers who were with the index case at the site and spent at least one night there in the previous 60 days. These individuals were contacted directly and enrolled at their preferred location, which could be their household, a participating health facility, or another private community setting (e.g., mosque or coffee shop). For both risk-based approaches, field teams aimed to conduct the response within three days of the case identification, which included traveling to the work site or other agreed location, administering a risk factor questionnaire on a tablet to assess potential risk factors for malaria, and collecting a dried blood sample (DBS) for microscopy and molecular analysis.

In addition to the risk-based RACD approaches described above, for all index cases with a local residence, household-based RACD (HH-RACD) was also conducted, according to provincial guidelines, by testing all household members and those in up to 25 neighboring households within 500 meters of the index case. Follow-up occurred within three days of case identification by a team of nurses and microscopists from district health facilities, who assessed treatment progress, administered the risk factor questionnaire on a tablet, and collected a DBS for microscopy and molecular analysis.

For all RACD approaches, malaria risk factor data collected included intervention use, forest and other travel and mobility patterns, history of malaria-related symptoms, residence household conditions, and socio-economic and demographic characteristics. Household– and individual-level questionnaires were developed in English then translated to Bahasa and Acehnese, then back translated by two fluent bilingual health experts before field testing. Data collection teams conducted community sensitization including notifying village leaders ahead of each RACD event. Up to two return visits were conducted in HH-RACD to maximize participation, and for VB-RACD, study teams would occasionally stay overnight at the venue/work site location because of long distances and difficult road conditions to reach the forest sites.

Microscopy or LAMP-positive individuals identified during RACD were notified immediately by health facility staff and an additional visit scheduled to collect a second blood sample (venous blood sample collected up to 3mL based on age), administer an additional questionnaire on a tablet, and provide treatment based on slide and LAMP results per national treatment guidelines.

### Laboratory procedures

Thick and thin blood smears were examined by trained microscopists at district health facilities (17). Smears were fixed and stained with 3% Giemsa. A thin smear was considered negative if no parasites were seen in 100 high-powered fields. If parasites were identified, an additional 100 high-powered fields were examined to determine species. An expert level-certified microscopist performed quality assurance at the Banda Aceh Provincial Hospital for all positive malaria cases and 10% of randomly selected negatives.

All DBS samples collected were dried overnight at the subdistrict health facilities then stored in plastic bags and sealed with desiccant before being transported to the Provincial Hospital in Banda Aceh for LAMP testing All DBS samples were stored at 4°C within one week of collection and then –20°C within one month of collection. Venous blood samples collected from positive cases were stored at 4°C for up to 5 days after collection. Once per quarter, blood samples were transported to Eijkman Institute in Jakarta for DNA extraction and then stored at –20 or –80°C. DNA from DBS was extracted using the Saponin/ Chelex method (18), while DNA from the EDTA/heparinized preserved blood was extracted using QiaAmp kits. Using 15 uL of Chelex extracted DNA, Pan-LAMP testing followed by *Pf*-LAMP-specific testing for Pan-LAMP positive samples was performed using a commercial Loopamp detection kit (19,20) in accordance with the manufacturer’s instructions (Eiken Chemical, Co., Ltd., Japan). All microscopy and Pan-LAMP positive samples and 10% of randomly selected LAMP negative samples from RACD were tested by the Eijkman Institute malaria laboratory using Chelex-extracted DNA from a second DBS. Nested cytochrome-B PCR testing targeting the 18S rRNA gene was used to identify the four human-only species of malaria (21), and *P. knowlesi* specific nPCR for all samples (22). All assays were internally validated using negative and low parasitemia positive controls.

### Data management and analysis

Field data collection was completed using an Open Data Kit (ODK) Collect v.1.5.1 form on Lenovo Tab 3 tablets with integrated geolocation. Data from tablets were uploaded to the study database in Eijkman Institute and UCSF servers for backup. All data were stored on password-protected computers and only accessible by study team members. Quantitative data were cleaned and analyzed using STATA (version 17) and R. The primary outcome measure was the yield of secondary cases identified through each RACD strategy.

### Ethical approval

This study was reviewed and approved by the Committee on Human Research at the University of California San Francisco (16–20220) and the Committee on Health Research Ethics of the National Institute of Health Research and Development, Indonesia Ministry of Health (LB.02.01/2/KE.083/2017). Written informed consent was obtained from all study participants. For individuals under the age of 18, informed consent and risk factor questionnaire was administered to a parent or guardian. For participants between the ages of 12-17, written informed assent was also obtained.

## RESULTS

### Index case enrollment

The study enrolled 47 index cases presenting to health facilities in the five target subdistricts (**Table 1** and **Figure 2**). Among these, 39 were initially diagnosed by microscopy at the health facility or district or provincial health office, and 8 were initially microscopy negative but tested positive by LAMP. Forty-six of the index cases were later confirmed by PCR (29 *P. knowlesi*, 17 *P. vivax*), while 1 index case that was initially diagnosed by microscopy was PCR negative. Forty-one of the index cases reported recent history of forest or forest-fringe work and 36 of these were eligible for peer-referral or venue-based response; four cases were from the same peer network or venue as another case, so a separate response was not conducted, and one case worked alone in the forest and had no eligible contacts. Six cases had no forest exposure and were eligible only for household response.

**Figure 2:**
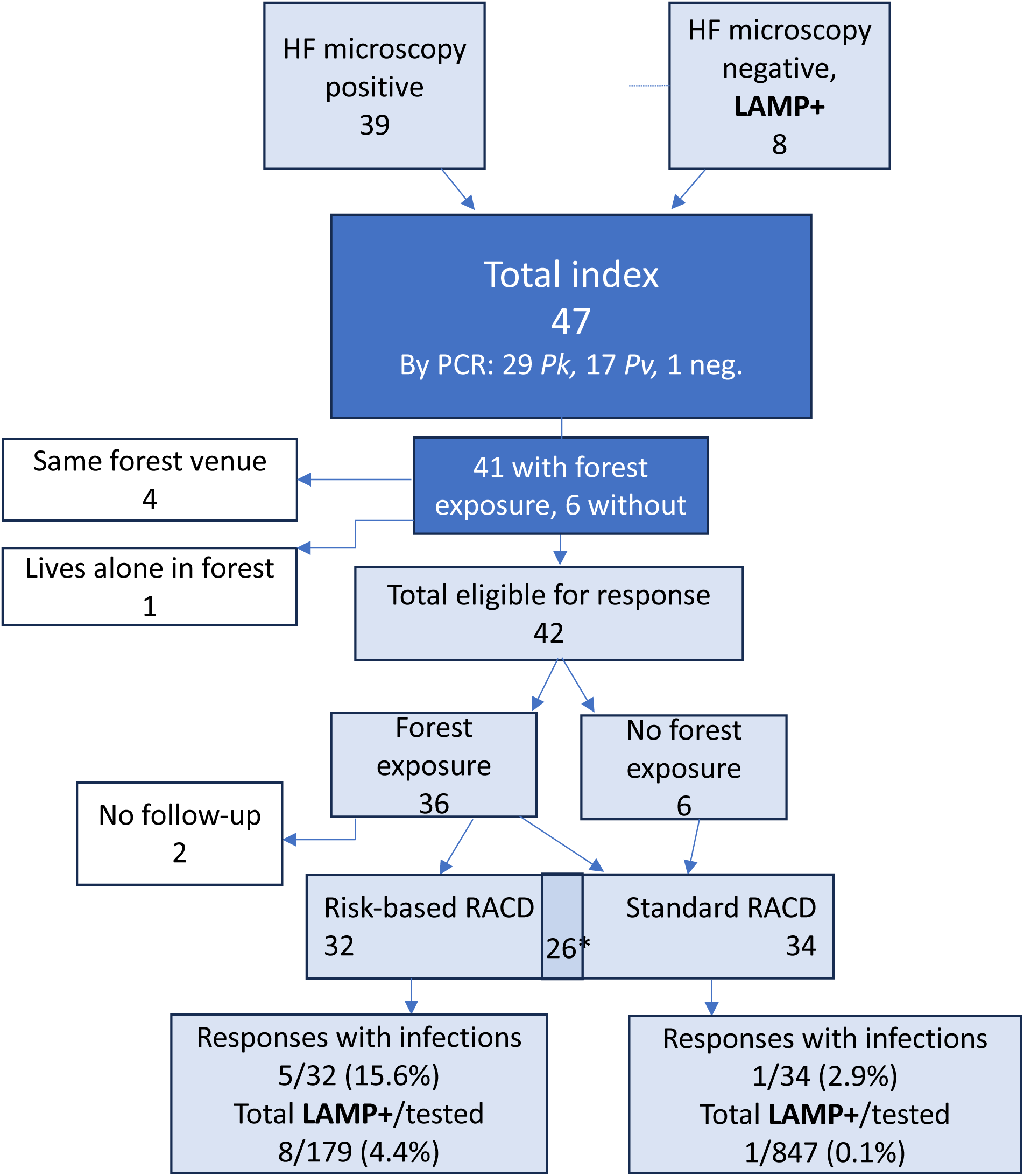
Index case enrollment and RACD response.

**Table 1:** Study recruitment and laboratory testing results.

| PCR-confirmed species | Index cases | RACD responses |  |  |  |
| --- | --- | --- | --- | --- | --- |
|  | Total (eligible and followed up) | PR<br>Eligible index cases (individuals tested) | VB<br>Eligible index cases (individuals tested) | Total Risk-based<br>Eligible index cases (individuals tested) | HH<br>Eligible index cases (individuals tested) |
| *P. knowlesi* | 24 (21) | 12 (30) | 6 (91) | 18 (121) | 18 (447) |
| *P. vivax* | 13 (11) | 4 (14) | 4 (20) | 8 (34) | 9 (221) |
| Undetermined | 5 (4) | 3 (12) | 1 (6) | 4 (18) | 4 (90) |
| Negative | 5 (4) | 1 (2) | 1 (4) | 2 (6) | 3 (89) |
| **Total** | 47 (40) | 20 (58) | 12 (121) | 32 (179) | 34 (847) |
| **LAMP +** | 9 (0.9%; 0.3-1.5%) | 5 (8.5%; 2.8 - 18.7%) | 3 (2.5%; 0.5-7.1%) | 8 (4.4%; 1.9-8.6%) | 1 (0.1%; 0.0-0.7%) |
| **LAMP/PCR +** | 4 (0.4%; 0.0-0.8%) | 0 (0.0%) | 3 (2.5%; 0.5-7.1%) | 3 (1.7%; 0.3-4.8%) | 1 (0.1%; 0.0-0.7%) |

Risk-based RACD was conducted for 32 of the index cases with forest exposure (20 peer-referral cases and 12 venue-based cases). Household-based RACD was conducted for 34 cases (26 of which also received PR or VB response).

### Yield of RACD-identified infections among different RACD strategies

From the 20 PR-RACD events conducted, 58 individuals provided consent and a blood sample for parasite testing, and from the 12 VB-RACD events conducted, 121 individuals provided consent and a blood sample, for an average of 2.9 and 10.1 individuals tested per PR-RACD and VB-RACD response, respectively. Of the total 32 risk-based RACD events, 5 identified additional infections (15.6%). Among 179 individuals tested during these events, 6 were microscopy positive (3 in VB-RACD and 3 in PR-RACD); all 6 were confirmed positive by LAMP, in addition to 2 microscopy negative individuals who were LAMP+, resulting in a LAMP positivity rate of 4.5% (95% CI 1.9-8.6%). However, only 3 were positive by PCR (all *P. vivax* in VB-RACD), for a positivity rate of 1.7% (95% CI 0.3-4.8%).

From the 34 HH-RACD events, a total of 881 out of 939 household members and neighbors who resided in the target screening area provided consent to participate in the study and 847 provided a blood sample (average of 24.9 individuals tested per event). Of the 34 HH-RACD events, only 1 identified an additional infection (2.9%). Of the 847 individuals tested during these events, 3 were microscopy positive, but only 1 was LAMP positive, for a positivity rate of 0.1% (95% CI 0.0-0.7%), and this was confirmed by PCR as *P. vivax*.

The yield for risk-based RACD was significantly higher than HH-RACD for both LAMP (p<.0001, Fisher’s exact test) and PCR (p=0.018). The overall refusal or absentee rate for RACD testing by approach was 12.1% for PR-RACD (8/66), 16.0% for VB-RACD (23/144), and 9.8% for household-based RACD 9.8% (92/939)

### Characteristics of the study population by index cases and RACD strategy

The vast majority of index cases were male (93.6%). (**Table 2**) Men also made up the majority of those screened by VB-RACD (94.2%) and PR-RACD (100%), but less than half by HH-RACD (43.5%). Logging, mining and other outdoor labor were the most common occupations among index cases (46.8%) and VB-RACD (68.6%) and PR-RACD (46.6%) participants, while farming was most common among HH-RACD (35.5%) participants.

**Table 2:** Characteristics of the study population by RACD screening strategy.

| **Variable** | **Index**<br>**n = 47** | **PR RACD**<br>**n = 58** | **VB RACD**<br>**n = 121** | **HH RACD**<br>**n = 881** |
| --- | --- | --- | --- | --- |
| **Age category** |  |  |  |  |
| *0-15* | 1 (2.1%) | 0 (0.0%) | 0 (0.0%) | 198 (22.5%) |
| *16-29* | 9 (19.1%) | 15 (25.9%) | 31 (25.6%) | 201 (22.8%) |
| *30-45* | 26 (55.3%) | 32 (55.2%) | 61 (50.4%) | 300 (34.1%) |
| *>45* | 11 (23.4%) | 11 (19%) | 29 (24%) | 182 (20.7%) |
| **Male** | 44 (93.6%) | 58 (100%) | 114 (94.2%) | 383 (43.5%) |
| **Some secondary education** | 27 (57.4%) | 39 (67.2%) | 72 (59.5%) | 458 (52.0%) |
| **Occupation** |  |  |  |  |
| *Farming* | 8 (17.0%) | 16 (27.6%) | 6 (5%) | 313 (35.5%) |
| *Logging, mining, outdoor labor* | 22 (46.8%) | 27 (46.6%) | 83 (68.6%) | 22 (2.5%) |
| *Student, teacher, housewife* | 0 (0.0%) | 1 (1.7%) | 0 (0.0%) | 275 (31.2%) |
| *Other* | 17 (36.2%) | 14 (24.1%) | 32 (26.4%) | 271 (30.8%) |
| **Subdistrict** |  |  |  |  |
| *Krueng Sabe* | 19 (40.4%) | 19 (32.8%) | 54 (44.6%) | 287 (32.6%) |
| *Kuta Cotgli* | 2 (4.3%) | 1 (1.7%) | 2 (1.7%) | 27 (3.1%) |
| *Lhoong* | 14 (29.8%) | 27 (46.6%) | 11 (9.1%) | 353 (40.1%) |
| *Sampoiniet* | 3 (6.4%) | 11 (19%) | 0 (0.0%) | 71 (8.1%) |
| *Saree* | 9 (19.1%) | 0 (0.0%) | 54 (44.6%) | 143 (16.2%) |
| **Slept under net the previous night** | 24 (51.1%) | 29 (50%) | 28 (23.1%) | 478 (54.9%) |
| **Main residence sprayed in past year** | 0 (0.0%) | 2 (3.4%) | 2 (1.7%) | 0 (0.0%) |
| **Main residence traditional roof or wall** | 20 (42.6%) | 20 (34.5%) | 30 (24.8%) | 65 (7.4%) |
| **Spent night in forest in past 60 days** | 40 (85.1%) | 52 (89.7%) | 44 (36.4%) | 40 (4.5%) |
| **Second residence in forest** | 30 (63.8%) | 30 (51.7%) | 59 (48.8%) | 29 (3.3%) |
| **Frequency of monkey sightings** |  |  |  |  |
| *None* | 20 (42.6%) | 35 (60.3%) | 50 (41.3%) | 543 (61.6%) |
| *At least every three days* | 11 (23.4%) | 8 (13.8%) | 10 (8.3%) | 218 (24.7%) |
| *Daily or every other day* | 16 (34.0%) | 15 (25.9%) | 61 (50.4%) | 120 (13.6%) |
| **Fever in the past 6 months** | 16 (34.0%) | 2 (3.4%) | 7 (5.8%) | 53 (6.0%) |

Index cases and those participating in VB-RACD and PR-RACD response were far more likely to have slept in the forest in the past 60 days (85.1%, 36.4%, and 89.2%, respectively), compared to HH-RACD (4.5%), as well as have a second residence in the forest (63.8%, 48.8%, and 51.7%, compared to 3.3%). Daily or every other day monkey sightings were also higher among index cases (34.0%), VB-RACD (50.4%), and PR-RACD (25.9%) compared to HH-RACD (13.6%).

## DISCUSSION

In the low-transmission, forest work-associated malaria setting of Aceh Province, Indonesia, targeting RACD to co-exposed contacts of malaria index cases substantially increased infection yield compared to standard household-based RACD. Our findings demonstrate that a targeted risk-based strategy is feasible to implement, and that individuals with shared occupational or travel exposures to index cases can be efficiently identified and tested for malaria infection. Although positivity rates were low overall, risk-based RACD using molecular detection identified more secondary infections per index case and achieved a higher positivity rate among those tested than household-based RACD.

Geographical clustering of malaria infections in lower transmission settings is well-documented, supporting the rationale for geographically targeted reactive response such as RACD (23). However, in low transmission settings in Southeast Asia and elsewhere where a predominant source of exposure is away from the home, geographically targeted reactive strategies will have less impact than reactive strategies targeting occupational or travel-related exposure (24). This is particularly relevant for *P. knowlesi*, where transmission depends on contact with forest-dwelling macaque reservoirs primarily through forest work (25). In our study, most index cases and their risk-based contacts worked in logging, mining, or other outdoor occupations and reported they spent a night in the forest or forest fringe in the past 60 days. Notably, a case-control study conducted contemporaneously with this study identified working and sleeping in the forest and having a second residence in the forest as key risk factors for infection (16). In contrast, household-based RACD participants were less likely to have recent forest exposure, which contributed to the markedly lower yield.

Our findings align with those from the Greater Mekong Subregion demonstrating improved yield of testing occupational contacts or co-travelers of index cases (6). Given the very low yield and relatively high cost of household-based RACD in these predominantly forest-area malaria settings (26), redirecting resources towards high-risk forest contacts of index cases may be both more effective and cost-efficient, even after accounting for the logistical and financial implications of traveling to forest-based worksites.

Diagnostic challenges remain a key constraint in very low malaria transmission settings. While microscopy remains the standard in most elimination programs, its detection limit (50-100 parasites/ul) misses many low-density, asymptomatic infections. In this study, health facility-based microscopists as well as provincial health-level microscopists read and provided quality assurance of all blood slides obtained during RACD, but parasite densities of the index cases ranged from less than 10 to more than 800 parasites per uL (unpublished). LAMP testing has been shown to improve infection and hotspot detection in low transmission settings (27) although in this very low transmission setting, discordance between LAMP and PCR results underscored the difficulty of detecting sub-patent parasitemia with molecular testing due to differing blood volumes and potential stochastic effects of very low parasite densities. Given these challenges, and the costs associated with molecular testing, presumptive treatment without testing is an option that warrants consideration, particularly in settings with forest workers (28)(29). However, administering an 8-aminoquinoline regime for radical cure of liver hypnozoites, critical for radical cure and elimination of *P. vivax* malaria, requires that all individuals should be tested for G6PD deficiency using a quantitative G6PD testing device prior to treatment. Importantly, evidence shows that G6PD testing can now be feasibly conducted at the community level by local health workers (30).

A main limitation of this study was the small sample size of index cases and low numbers of RACD-identified infections detected, which is a common challenge for studies carried out in very low transmission settings. Relatedly, the discordant results between LAMP and PCR results for RACD participants made direct positivity comparisons between approaches more difficult. Third, under-enrollment could have occurred if index cases withheld peer contact or venue-related information due to sensitive or illegal forest-based activities. Finally, although our study had a quick turnaround time from index case presentation to RACD follow-up, we did not assess whether delays in receiving LAMP results for initially microscopy-negative cases affected the timeliness and completeness of follow-up.

Despite these challenges, this study was embedded within existing provincial and district malaria program activities, enhancing its operational relevance. Findings add to the limited operational evidence base supporting risk-based RACD strategies in predominantly *P. knowlesi*-endemic settings, and illustrates how molecular methods can be used programmatically to identify additional infections when conducting RACD.

## CONCLUSIONS

In low malaria transmission forest– and forest-fringe areas, targeting RACD by shared occupational and travel risk factors may lead to an increased detection of infections. Risk-based RACD was able to effectively identify individuals who recently had contact with an index case and identified more confirmed and probable infections compared to household-based RACD. Although additional operational logistics and costs should be considered, risk-based RACD can be an effective component of a malaria elimination strategy in forest-going populations.

## Data Availability

All de-identified data produced in this study are available upon reasonable request to the authors.

## Acknowledgements

The authors appreciate the patients and study participants who contributed blood samples and survey responses to the study. The authors are grateful to the malaria program staff of the National Malaria Control Program, the Aceh Province Health Office, Aceh Besar and Aceh Jaya District Health Offices, and the nurses and microscopists in the subdistricts of Kuta Cot Gli, Lembah Seulawah, Lhoong (Aceh Besar district) and in Krueng Sabe and Sampoiniet (Aceh Jaya district) for their valuable contributions and support in study implementation.

